# Reimbursement for Maxillofacial Surgery: Regional Analysis of the Physician Fee Schedule Reveals Unique Payment Increases in the Western United States From 2010 to 2019

**DOI:** 10.1101/2023.03.15.23287342

**Authors:** Angelina Thuy Tran, Michael Joseph Diaz, Sai Batchu

**Author notes:** **Financial Disclosure:** None. **IRB Approval:** Exempt. **Patient Consent:** N/A.

## Abstract

**Introduction:** Medicare Part B reimbursement data has been thoroughly evaluated in several surgical subspecialties to date, with significant ramifications for continued policy research and payment reform discussions. However, trends in maxillofacial surgery payment remain unstudied.

**Methods:** In this cross-sectional study of Part B reimbursement data, we analyzed regional trends in reimbursement for common maxillofacial repair and reconstruction procedures (N=17). All payment data converted to January 2019 United States dollars.

**Results:** From 2010 to 2019, fees for maxillofacial surgery evidenced strong region- and procedures-specific changes. After adjusting for inflation, the Southern United States reported an unweighted mean change in Medicare reimbursement of negative 12.59% (SD = 2.63%), while the Western United States reported an unweighted mean change in Medicare reimbursement of +0.08% (SD = 2.82%). In the Midwest, only Part B billing for reconstruction of the mandibular rami and/or body without internal rigid fixation (CPT code 21195) kept up with inflation. In the West, 8 CPT codes (47%) kept up with inflation: 21422, 21246, 21196, 21195, 21194, 21193, 21048, and 21046. In the Northeast and the South, all analyzed procedures reported decreased reimbursement rates after adjusting for inflation.

**Conclusion:** Reimbursement for maxillofacial surgery has lagged behind inflation with pronounced regional bias.

## Introduction

Maxillofacial repair and reconstruction (MRR) broadly involve surgical correction of facial lacerations, jaw/facial bone fractures, and tumor/cyst removal requiring osteotomy.^1-3^ These procedures can restore the patient’s facial appearance, chewing function, and overall quality of life. MRR procedures commonly involve bone grafting, dental implant placement, and soft tissue reconstruction, among other techniques.^4,5^ The primary goal is to restore function and aesthetics while minimizing scarring and optimizing healing.

Most commonly, these procedures are performed by oral and maxillofacial (OMF) surgeons in the emergency setting. Indeed, even amongst preprofessional students, it is generally well understood that OMF surgeons are to be consulted in the event of facial trauma, dento-facial deformities, mandibular reconstruction and temporomandibular joint surgery.^6^ For these specialists, reimbursement rates have decreased markedly, with acceleration seen in recent years.^7^ It should also be noted that the ongoing provider payment debate (and supportive evidence) is not unique to twenty-first century surgery. As far back as the early 1990s, Reagan et al. reported survey results revealing that dental procedures in the hospital setting were unequally reimbursed.^8^ Given the importance of these procedures for quality of life and related implications of Medicare reimbursement rates on the availability and accessibility of maxillofacial surgery, there is a need to understand the changing fee landscape.

Despite efforts to account for regional differences in costs of living, wages, and other demographic and health factors in procedural fee determination (i.e., the geographic practice cost index), geographic variation in Medicare spending remains explicit.^9,10^ Thus, the objective of the current study is to identify geographic payment trends for MRR surgeries covered by Medicare. This information can be used to inform policy decisions aimed at improving access to quality care while ensuring the financial sustainability of Medicare programs.

## Methods

### Data Retrieval

Part B pricing information for common MRR Current Procedural Terminology (CPT) codes was obtained from the Medicare Physician Fee Schedules (MPFS) datasets. At the time of writing, the MPFS additionally defines dentist reimbursement for Medicare-eligible dental services. Payment data for years 2010 and 2019 were retrieved. See Table 1 for descriptions and corresponding CPT codes for all included MRR procedures (N=17). Of note, “unlisted craniofacial and maxillofacial procedure” (CPT code 21299) was not included in the present analysis due to unavailable 2010 MPFS data.

**Table 1.** Summary of CPT codes for common maxillofacial repair and reconstruction procedures.

| **CPT Code\*** | **Description** | **2019 National Payment Amount** |
| --- | --- | --- |
| **21470** | Open treatment of complicated mandibular fracture by multiple surgical approaches including internal fixation, interdental fixation, and/or wiring of dentures or splints | \$1,244.79 |
| **21465** | Open treatment of mandibular condylar fracture | \$933.77 |
| **21454** | Open treatment of mandibular fracture with external fixation | \$565.45 |
| **21423** | Open treatment of palatal or maxillary fracture (LeFort I type); complicated (comminuted or involving cranial nerve foramina), multiple approaches | \$800.79 |
| **21422** | Open treatment of palatal or maxillary fracture (LeFort I type) | \$683.66 |
| **21247** | Reconstruction of mandibular condyle with bone and cartilage autografts (includes obtaining grafts) (e.g., for hemifacial microsomia) | \$1,705.73 |
| **21246** | Reconstruction of mandible or maxilla, subperiosteal implant; complete | \$916.84 |
| **21244** | Reconstruction of mandible, extraoral, with transosteal bone plate (e.g., mandibular staple bone plate) | \$1,081.89 |
| **21196** | Reconstruction of mandibular rami and/or body, sagittal split; with internal rigid fixation | \$1,520.13 |
| **21195** | Reconstruction of mandibular rami and/or body, sagittal split; without internal rigid fixation | \$1,477.96 |
| **21194** | Reconstruction of mandibular rami, horizontal, vertical, C, or L osteotomy; with bone graft (includes obtaining graft) | \$1,527.70 |
| **21193** | Reconstruction of mandibular rami, horizontal, vertical, C, or L osteotomy; without bone graft | \$1,329.48 |
| **21049** | Excision of benign tumor or cyst of maxilla; requiring extra-oral osteotomy and partial maxillectomy (e.g., locally aggressive or destructive lesion(s)) | \$1,250.92 |
| **21048** | Excision of benign tumor or cyst of maxilla; requiring intra-oral osteotomy (e.g., locally aggressive or destructive lesion(s)) | \$1,155.77 |
| **21047** | Excision of benign tumor or cyst of mandible; requiring extra-oral osteotomy and partial mandibulectomy (e.g., locally aggressive or destructive lesion(s)) | \$1,358.31 |
| **21046** | Excision of benign tumor or cyst of mandible; requiring intra-oral osteotomy (e.g., locally aggressive or destructive lesion(s)) | \$1,137.75 |
| **21045** | Excision of malignant tumor of mandible; radical resection | \$1,265.69 |

\*CPT code 21299 (“UNLISTED CRANIOFACIAL AND MAXILLOFACIAL PROCEDURE”) not included in the present analysis by reason of unavailable MPFS data.

### Analysis

Payment localities were geographically mapped to the four United States (U.S.) regions (Northeast, Midwest, South, and West) as designated by the U.S. Census Bureau. A full tabulation of included procedure codes in provided in Table 1. Payment data was converted to January 2019 U.S. dollars using U.S. Bureau of Labor Statistics consumer price index inflation data. Descriptive statistics were generated using programming language R v4.2.2.

## Results

### Northeast Fee Trends

MRR reimbursement rates in the Northeastern United States reported a mean inflation-adjusted change of negative 8.37% (SD = 2.69%, unweighted) between years 2010 and 2019. All MRR procedures observed a decrease in reimbursement. Excision of benign tumor or cyst of maxilla requiring extra-oral osteotomy and partial maxillectomy (CPT code 21049) reported the greatest inflation-adjusted change in reimbursement (−12.84%) during this time (1508 USD to 1315 USD, CAGR: -0.015). Reconstruction of mandibular rami and/or body without internal rigid fixation (CPT code 21195) reported the lowest inflation-adjusted change in reimbursement (−3.27%) over this period (1614 USD to 1561 USD, CAGR: -0.004). Additional granular data is reported in Table 2.

**Table 2.** Maxillofacial surgery reimbursement rate trends in the **Northeastern region** of the United States, based on 2010-2019 Medicare Physician Fee Schedule data.

| **CPT Code** | **Average Reimbursement Rate in 2010 (Unadjusted)** | **Average Reimbursement Rate in 2010 (Jan 2019 USD)** | **Average Reimbursement Rate in 2019** | **Adjusted Change in Reimbursement (2010-2019) (%)** | **CAGR** |
| --- | --- | --- | --- | --- | --- |
| **21470** | \$1238.72 | \$1436.91 | \$1312.09 | -8.69 | -0.01005 |
| **21465** | \$962.34 | \$1116.31 | \$984.46 | -11.81 | -0.01387 |
| **21454** | \$571.68 | \$663.14 | \$597.11 | -9.96 | -0.01159 |
| **21423** | \$829.03 | \$961.67 | \$844.65 | -12.17 | -0.01431 |
| **21422** | \$685.06 | \$794.67 | \$722.04 | -9.14 | -0.01059 |
| **21247** | \$1690.38 | \$1960.84 | \$1797.66 | -8.32 | -0.00961 |
| **21246** | \$871.21 | \$1010.61 | \$966.55 | -4.36 | -0.00494 |
| **21244** | \$1091.75 | \$1266.43 | \$1142.80 | -9.76 | -0.01135 |
| **21196** | \$1509.78 | \$1751.34 | \$1603.06 | -8.47 | -0.00978 |
| **21195** | \$1390.99 | \$1613.55 | \$1560.87 | -3.27 | -0.00368 |
| **21194** | \$1457.65 | \$1690.88 | \$1610.05 | -4.78 | -0.00543 |
| **21193** | \$1283.55 | \$1488.91 | \$1401.31 | -5.88 | -0.00671 |
| **21049** | \$1300.40 | \$1508.46 | \$1314.78 | -12.84 | -0.01515 |
| **21048** | \$1139.12 | \$1321.38 | \$1220.95 | -7.60 | -0.00874 |
| **21047** | \$1342.55 | \$1557.36 | \$1430.00 | -8.18 | -0.00943 |
| **21046** | \$1118.79 | \$1297.80 | \$1202.40 | -7.35 | -0.00845 |
| **21045** | \$1273.31 | \$1477.04 | \$1332.84 | -9.76 | -0.01135 |

### Midwest Fee Trends

MRR reimbursement rates in the Midwestern United States reported a mean inflation-adjusted change of negative 4.47% (SD = 2.57%, unweighted) between years 2010 and 2019. Excision of benign tumor or cyst of maxilla requiring extra-oral osteotomy and partial maxillectomy (CPT code 21049) reported the greatest inflation-adjusted change in reimbursement (−9.22%) during this time (1347 USD to 1223 USD, CAGR: -0.011). Reconstruction of mandibular rami and/or body without internal rigid fixation (CPT code 21195) reported a positive inflation-adjusted change in reimbursement (+0.44%) over this period (1433 USD to 1439 USD, CAGR: 0.0005). Additional granular data is reported in Table 3.

**Table 3.** Maxillofacial surgery reimbursement rate trends in the **Midwestern region** of the United States, based on 2010-2019 Medicare Physician Fee Schedule data.

| **CPT Code** | **Average Reimbursement Rate in 2010 (Unadjusted)** | **Average Reimbursement Rate in 2010 (Jan 2019 USD)** | **Average Reimbursement Rate in 2019** | **Adjusted Change in Reimbursement (2010-2019) (%)** | **CAGR** |
| --- | --- | --- | --- | --- | --- |
| **21470** | \$1098.61 | \$1274.39 | \$1214.26 | -4.72 | -0.00536 |
| **21465** | \$848.78 | \$984.59 | \$910.76 | -7.50 | -0.00862 |
| **21454** | \$505.21 | \$586.04 | \$550.62 | -6.04 | -0.00690 |
| **21423** | \$729.18 | \$845.85 | \$780.54 | -7.72 | -0.00889 |
| **21422** | \$604.79 | \$701.55 | \$665.53 | -5.13 | -0.00584 |
| **21247** | \$1496.79 | \$1736.27 | \$1664.31 | -4.14 | -0.00469 |
| **21246** | \$778.13 | \$902.63 | \$894.29 | -0.92 | -0.00103 |
| **21244** | \$961.44 | \$1115.28 | \$1052.96 | -5.59 | -0.00637 |
| **21196** | \$1340.78 | \$1555.31 | \$1482.03 | -4.71 | -0.00535 |
| **21195** | \$1235.16 | \$1432.79 | \$1439.05 | 0.44 | 0.00048 |
| **21194** | \$1302.79 | \$1511.24 | \$1490.58 | -1.37 | -0.00153 |
| **21193** | \$1136.61 | \$1318.47 | \$1297.04 | -1.63 | -0.00182 |
| **21049** | \$1161.63 | \$1347.49 | \$1223.24 | -9.22 | -0.01069 |
| **21048** | \$1006.66 | \$1167.72 | \$1124.96 | -3.66 | -0.00414 |
| **21047** | \$1199.74 | \$1391.70 | \$1326.52 | -4.68 | -0.00532 |
| **21046** | \$987.55 | \$1145.56 | \$1106.99 | -3.37 | -0.00380 |
| **21045** | \$1133.78 | \$1315.18 | \$1235.64 | -6.05 | -0.00691 |

### South Fee Trends

MRR reimbursement rates in the Southern United States reported a mean inflation-adjusted change of negative 12.59% (SD = 2.63%, unweighted) between years 2010 and 2019. Complicated open treatment of palatal or maxillary fracture (LeFort I type) (CPT code 21423) reported the greatest inflation-adjusted change in reimbursement (−16.78%) during this time (950 USD to 790 USD, CAGR: -0.020). Reconstruction of mandibular rami and/or body without internal rigid fixation (CPT code 21195) reported the lowest inflation-adjusted change in reimbursement (−7.51%) over this period (1576 USD to 1458 USD, CAGR: -0.009). Additional granular data is reported in Table 4.

**Table 4.** Maxillofacial surgery reimbursement rate trends in the **Southern region** of the United States, based on 2010-2019 Medicare Physician Fee Schedule data.

| **CPT Code** | **Average Reimbursement Rate in 2010 (Unadjusted)** | **Average Reimbursement Rate in 2010 (Jan 2019 USD)** | **Average Reimbursement Rate in 2019** | **Adjusted Change in Reimbursement (2010-2019) (%)** | **CAGR** |
| --- | --- | --- | --- | --- | --- |
| **21470** | \$1221.97 | \$1417.49 | \$1229.91 | -13.23 | -0.01565 |
| **21465** | \$953.26 | \$1105.78 | \$922.61 | -16.56 | -0.01992 |
| **21454** | \$555.98 | \$644.93 | \$557.74 | -13.52 | -0.01601 |
| **21423** | \$818.78 | \$949.79 | \$790.44 | -16.78 | -0.02020 |
| **21422** | \$666.35 | \$772.97 | \$673.98 | -12.81 | -0.01511 |
| **21247** | \$1681.67 | \$1950.73 | \$1686.00 | -13.57 | -0.01607 |
| **21246** | \$854.79 | \$991.56 | \$905.94 | -8.63 | -0.00998 |
| **21244** | \$1063.29 | \$1233.41 | \$1066.20 | -13.56 | -0.01606 |
| **21196** | \$1474.91 | \$1710.90 | \$1500.90 | -12.27 | -0.01444 |
| **21195** | \$1358.68 | \$1576.07 | \$1457.69 | -7.51 | -0.00864 |
| **21194** | \$1431.39 | \$1660.41 | \$1510.02 | -9.06 | -0.01049 |
| **21193** | \$1283.69 | \$1489.08 | \$1313.96 | -11.76 | -0.01380 |
| **21049** | \$1276.05 | \$1480.22 | \$1238.31 | -16.34 | -0.01963 |
| **21048** | \$1108.67 | \$1286.05 | \$1139.43 | -11.40 | -0.01336 |
| **21047** | \$1318.35 | \$1529.29 | \$1343.51 | -12.15 | -0.01429 |
| **21046** | \$1087.81 | \$1261.86 | \$1121.24 | -11.14 | -0.01304 |
| **21045** | \$1249.83 | \$1449.81 | \$1251.27 | -13.69 | -0.01623 |

### West Fee Trends

MRR reimbursement rates in the Western United States reported a mean inflation-adjusted change of 0.08% (SD = 2.82%, unweighted) between years 2010 and 2019. Of the 17 MRR procedures, 8 (47%) observed an increase in mean inflation-adjusted reimbursement in the Western United States. Reconstruction of mandibular rami and/or body without internal rigid fixation (CPT code 21195) reported the most positive, inflation-adjusted change in reimbursement (+5.80%) over this period (1450 USD to 1534 USD, CAGR: +006). Excision of benign tumor or cyst of maxilla requiring extra-oral osteotomy and partial maxillectomy (CPT code 21049) reported the most negative, inflation-adjusted change in reimbursement (−5.43%) over this period (1364 USD to 1290 USD, CAGR: -0.006). Additional granular data is reported in Table 5.

**Table 5.** Maxillofacial surgery reimbursement rate trends in the **Western region** of the United States, based on 2010-2019 Medicare Physician Fee Schedule data.

| **CPT**<br>**Code** | **Average**<br>**Reimbursement**<br>**Rate in 2010**<br>**(Unadjusted)** | **Average**<br>**Reimbursement**<br>**Rate in 2010**<br>**(Jan 2019 USD)** | **Average**<br>**Reimbursement**<br>**Rate in 2019** | **Adjusted**<br>**Change in**<br>**Reimbursement**<br>**(2010-2019) (%)** | **CAGR** |
| --- | --- | --- | --- | --- | --- |
| **21470** | \$1114.96 | \$1293.36 | \$1285.84 | -0.58 | -0.00065 |
| **21465** | \$863.21 | \$1001.33 | \$963.81 | -3.75 | -0.00423 |
| **21454** | \$510.79 | \$592.52 | \$586.68 | -0.99 | -0.00110 |
| **21423** | \$741.22 | \$859.81 | \$830.40 | -3.42 | -0.00386 |
| **21422** | \$611.59 | \$709.44 | \$711.33 | 0.27 | 0.00029 |
| **21247** | \$1523.35 | \$1767.09 | \$1758.55 | -0.48 | -0.00054 |
| **21246** | \$787.61 | \$913.63 | \$946.07 | 3.55 | 0.00388 |
| **21244** | \$972.98 | \$1128.66 | \$1127.59 | -0.09 | -0.00011 |
| **21196** | \$1356.46 | \$1573.50 | \$1574.73 | 0.08 | 0.00009 |
| **21195** | \$1249.58 | \$1449.51 | \$1533.58 | 5.80 | 0.00628 |
| **21194** | \$1318.86 | \$1529.88 | \$1574.99 | 2.95 | 0.00323 |
| **21193** | \$1158.65 | \$1344.04 | \$1370.99 | 2.01 | 0.00221 |
| **21049** | \$1175.80 | \$1363.93 | \$1289.87 | -5.43 | -0.00618 |
| **21048** | \$1018.01 | \$1180.89 | \$1201.31 | 1.73 | 0.00191 |
| **21047** | \$1214.57 | \$1408.90 | \$1399.76 | -0.65 | -0.00072 |
| **21046** | \$998.56 | \$1158.33 | \$1183.75 | 2.19 | 0.00242 |
| **21045** | \$1148.25 | \$1331.97 | \$1307.32 | -1.85 | -0.00207 |

## Discussion

In the present study, we analyzed Medicare payment trends in maxillofacial repair and reconstruction surgery over the 2010s. A primary motivation for studying these trends are their significant impact on the financial viability of private practices and, relatedly, access to care for Medicare beneficiaries. After adjusting for CPI inflation, we found that the Southern United States observed the greatest decline in MRR reimbursement, whereas the Western United States reported positive growth in MRR reimbursement. All procedural codes reported a net-negative change in reimbursement, corroborating prior evidence of national (yet variable) decreases in Medicare reimbursement for common facial trauma procedures.^7,11^

Geographic differences in Medicare reimbursement can have significant implications for healthcare providers, patients, and insurance companies. For providers, lower reimbursement may realistically limit investment in new equipment and may also discourage healthcare providers from practicing in areas with lower reimbursement rates, limiting access to care for patients in those regions. On the other hand, higher rates of reimbursement in certain areas may attract more providers to a fault, leading to an oversupply of healthcare services that are not necessarily needed, driving up overall costs. For patients, geographic variation in reimbursement rates can cause disparities in access to care and quality of service. Patients in areas with lower reimbursement rates may struggle to find healthcare providers, especially specialists, as they may move to areas where reimbursement rates are higher.^12^ Additionally, lower reimbursement rates may result in reduced services or fewer treatment options for patients, leading to lower-quality care.^13^ More research is needed to determine the associative strength of surgeon reimbursement with patient health outcomes.

Limitations of the present analysis include the lack of state- and locality-level granularity, year-over-year change statistics, and missed opportunity for weighting with quality procedural utilization data. Nonetheless, the finding shared here put forth valuable talking points towards ongoing Medicare payment reform discussions.

## Data Availability

All data is publicly available at CMS.gov.

## Conclusion

Geographic differences in Medicare reimbursement have complex and far-reaching implications for healthcare providers, patients, and insurance companies. In the present study, we reported first efforts to quantify longitudinal trends in Part B reimbursement for established maxillofacial repair and reconstruction procedures. Our approach identified a wealth of striking regional differences in reimbursement for maxillofacial surgery. Addressing these disparities is crucial to ensuring equitable access to high-quality healthcare for all Americans.

## Acknowledgements

None

